# Understanding the chronic kidney disease landscape using patient representation learning from electronic health records

**DOI:** 10.1101/2022.10.25.22280440

**Authors:** Karen Kapur, Moritz Freidank, Michael Rebhan

## Abstract

Understanding various subpopulations in chronic kidney disease can improve patient care and aid in developing treatments targeted to patients’ needs. Due to the general slow disease progression, electronic health records, which comprise a rich source of longitudinal real-world patient-level information, offer an approach for generating insights into disease. Here we apply the open-source ConvAE framework to train an unsupervised deep learning network using a real-world kidney disease cohort consisting of 2.2 million US patients from the OPTUM® EHR database. Numerical patient representations derived from ConvAE are used to derive disease subtypes, inform comorbidities and understand rare disease populations. To identify patients at high risk to develop end-stage kidney disease, we extend a validated algorithm classifying disease severity to hypothesize subpopulations of rapid chronic kidney disease progressors. We demonstrate that using a combination of data-driven methods offers a powerful exploratory approach to understand disease heterogeneity and identify high-risk patients who could be targeted for early therapeutic intervention to prevent end-stage kidney disease.

## Introduction

Electronic health records (EHR) comprise a rich source of longitudinal real-world patient-level information, consisting of a variety of different data types, including demographics, medical history, medications, laboratory test results and free-text clinical notes. There are many examples analyzing EHR data to generate knowledge about diseases (Chen and Sarkar 2014). In particular, large-scale population-based studies have been instrumental to uncover relationships between diseases (Dong et al. 2021) and to understand disease progression patterns (Jensen et al. 2014; Westergaard et al. 2019).

Using EHRs for clinical research poses several analytic challenges (Wei and Denny 2015, Goldstein 2020), as the data were collected for routine clinical care and were not designed for research purposes. To identify an accurate set of patients with a given condition, one can employ electronic phenotyping which uses sophisticated algorithms to label which patients have a given medical condition. Phenotyping algorithms have been developed for a variety of diseases and many definitions can be found in the Phenotype KnowledgeBase (PheKB) (Kirby et al. 2016). However, electronic phenotyping is time consuming and requires extensive collaboration with experts having domain knowledge.

Recent data-driven approaches have proposed a more scalable approach to defining homogeneous patient subpopulations. Longitudinal patient health information, as captured in EHRs, has been used by a variety of statistical or machine learning approaches to identify groups with similar patient patterns (Basile and Ritchie 2018; Allam et al. 2021). EHR phenotyping can be considered as an unsupervised machine learning problem to cluster similar patients (Drysdale et al. 2017; Fereshtehnejad et al. 2017; Zhang et al. 2019; Z. Xu et al. 2020). A subset of such methods employs patient representation learning, which transforms high-dimensional sequences of medical events into a numerical representation (Si et al. 2021). A method employing a convolutional autoencoder (ConvAE) recently demonstrated the ability to derive disease sub-types from EHRs at scale (Landi et al. 2020a). Using patients’ sequences of historical medical codes, the method identifies groups of similar patients, representing disease subtypes. The disease subtypes were shown to be clinically meaningful and interpretable in terms of comorbidities, disease progression and symptom severity.

In this paper we apply data-driven approaches including both machine-learning methods deriving patient representations and electronic phenotyping algorithms to understand a chronic kidney disease (CKD) population. CKD is a chronic disease with significant unmet medical need (Torreggiani et al. 2021) and comprises a heterogeneous set of disorders, with multiple factors contributing to the disease, various comorbidities (Shafi and Coresh 2019), as well as variable disease progression rates (Go et al. 2018; Tsai et al. 2017; Remuzzi, Benigni, and Remuzzi 2006). We apply the open-source ConvAE framework (Landi et al. 2020a) to train an unsupervised deep learning architecture using a real-world kidney disease cohort consisting of 2.2 million US patients from OPTUM® EHR database, which consists of observational data obtained from a variety of healthcare providers, including primary and secondary care. We demonstrate the utility of these patient representations to derive CKD subtypes, inform comorbidities and understand rare disease populations. We extend a validated CKD staging electronic phenotype (Shang et al. 2021) to derive CKD progression labels which we correlate with patient representations to derive hypotheses of rapid CKD progressor sub-types. Applying patient representation learning to a large observational CKD cohort offers a powerful exploratory approach to understand disease and hypothesize high risk patient sub-populations, with the aim of improving patient care and developing treatments targeted to patients’ needs. Due to the limited capacity for kidney regeneration(Yang, Liu, and Fogo 2014), identification of high-risk CKD patients who could be targeted for early therapeutic intervention is vital to prevent end-stage kidney disease.

## Results

A cohort of 2.2 million patients was extracted from the OPTUM® EHR database, a US-based patient population, with selection criteria detailed in Table 1 and patient characteristics described in Table 2. Patients were selected to have at least one kidney disease related diagnosis or procedure code as well as a minimal follow-up duration of 2 years and multiple serum creatinine measurements from which estimated glomerular filtration rate (eGFR) was calculated. The resulting population consisted of patients with an average length of follow-up of 10 years, an average of 29 serum creatinine measurements, roughly balanced by gender and enriched for Caucasian race.

**Table 1:** Cohort selection criteria applied to Optum EHR. Kidney disease diagnosis/procedure codes are listed in Supplementary Table 1

| Criteria | Number of patients |
| --- | --- |
| All patients | 104,114,518 |
| IDN indicator = 1 | 87,669,592 |
| Kidney disease diagnosis/procedure codes | 6,243,647 |
| ≥ 2 year follow-up<br>≥ 15 serum creatinine measurements | 2,219,789 |

**Table 2:**
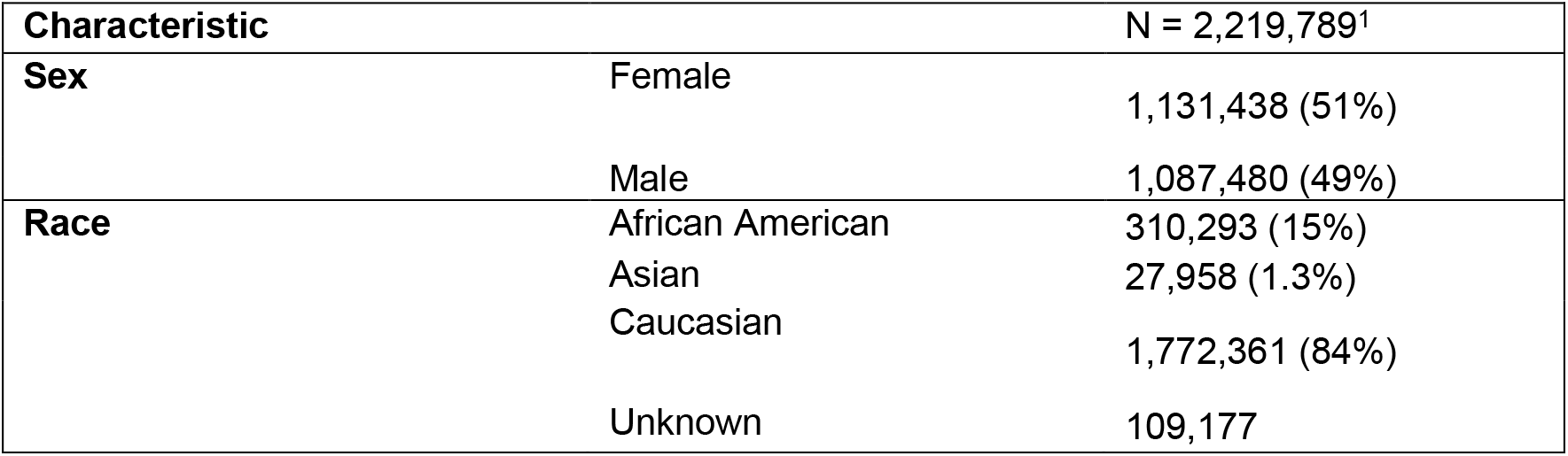
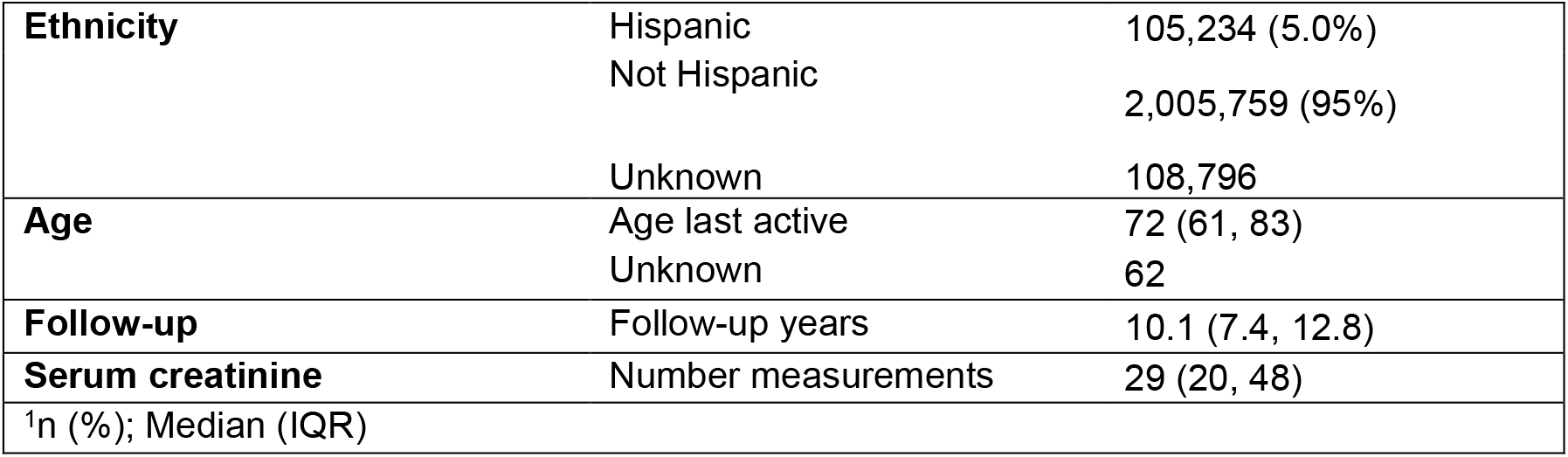
Characteristics of the kidney disease cohort.

We fit the ConvAE (Landi et al. 2020b) model using patients’ sequence of diagnosis and procedure codes. For each patient, the method yields a 100-dimensional numerical vector, or embedding. Using k-means clustering, we clustered patients, determining the optimal number of clusters using the elbow method (Thorndike 1953), identifying 5 clusters (Supplementary Figure 1). We visualize the different patients and their clusters using a uniform manifold approximation and projection for dimension reduction (UMAP) (McInnes, Healy, and Melville 2018) projection of the patient-level embeddings into two dimensions, as shown in Figure 1.

**Figure 1:**
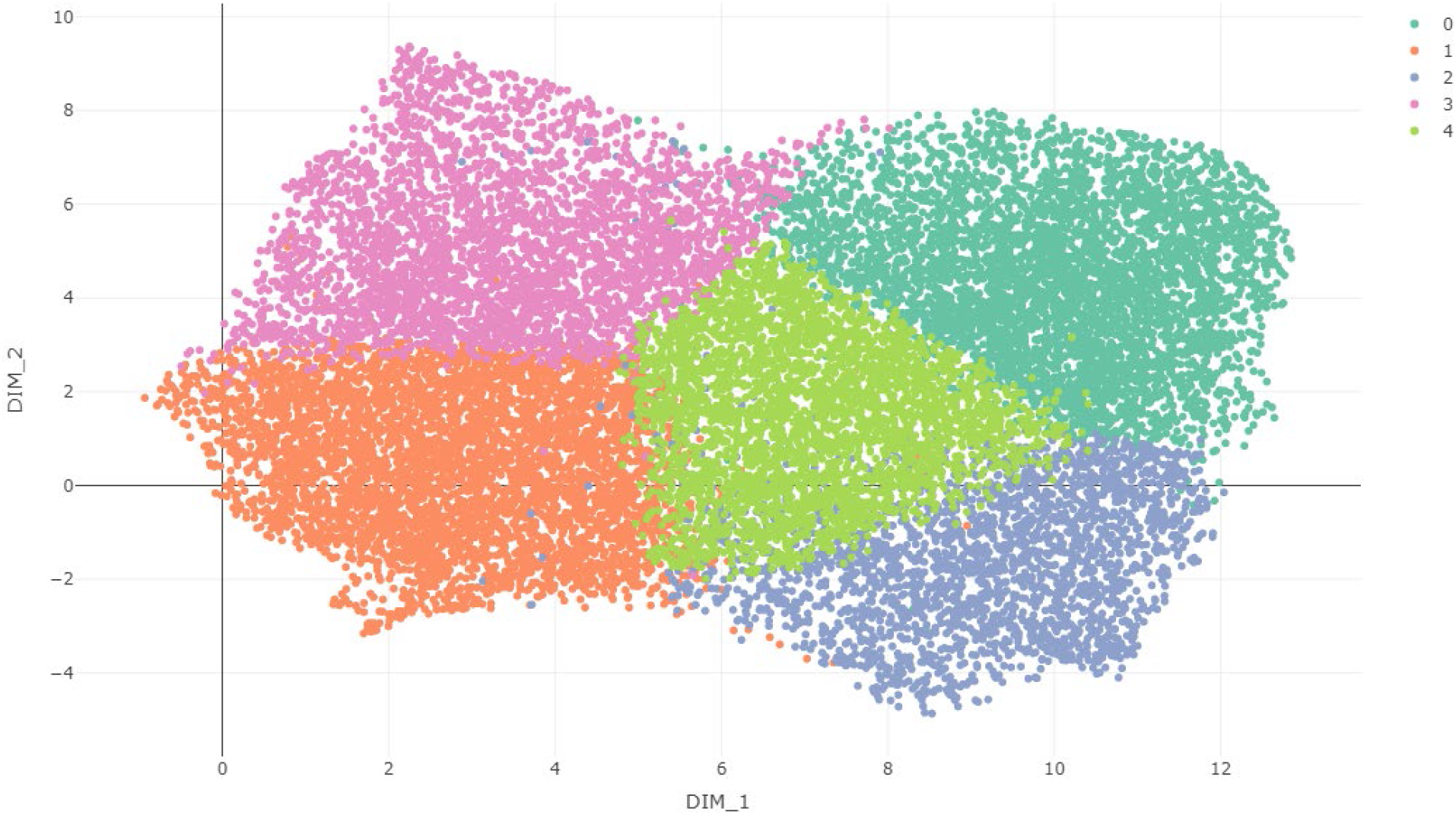
UMAP projection of patient-level embeddings colored by cluster membership for K=5 clusters. A random 1% of patients are plotted to avoid over-plotting.

To interpret each cluster, we selected the top over- and under-enriched terms and compare the term frequencies in each cluster, as shown in Figure 2. Briefly, Group 0 is enriched for outpatient visits, immunizations and screenings, whereas Group 1 is enriched for acute events, including acute renal failure and sepsis. Group 2 is enriched for chemotherapy as well as surgical pathology. Group 3 is enriched for heart disease codes including congestive heart failure and atrial fibrillation. Group 4 is enriched for emergency department visits, infusions, pain, anxiety and injections. A full list of term frequencies and descriptions can be found in Supplementary Table 2. Applying the ConvAE method to generate patient-level embeddings, we generate visual and interpretable summaries that aid in understanding different facets of this CKD disease population.

**Figure 2:**
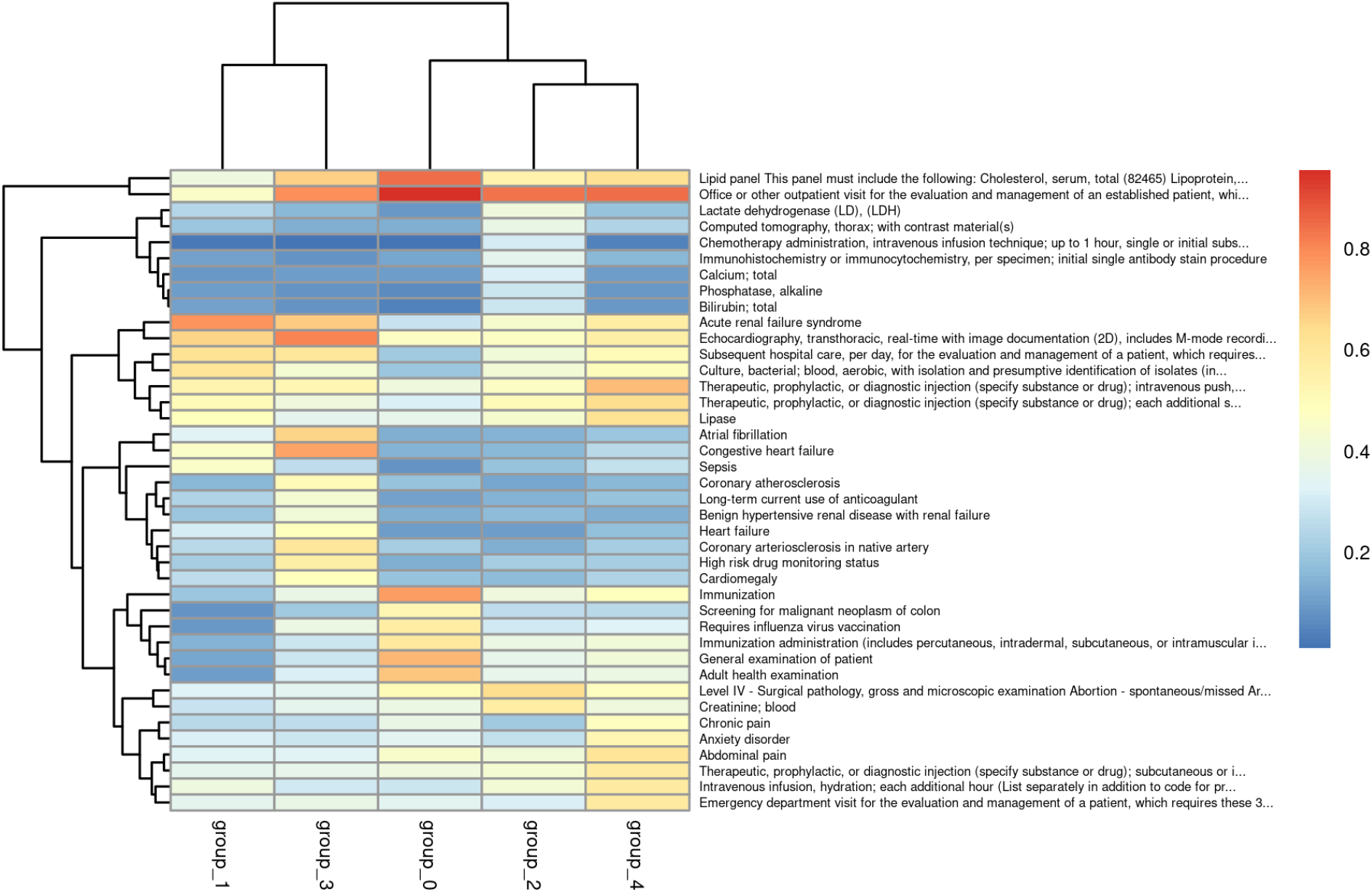
Heatmap displaying the proportion of patients with a given code for each of the K=5 clusters. The top 10 codes with largest absolute differences are shown here. A full list can be found in Supplementary Table 2.

We further overlaid annotations on the UMAP plots, to yield insights into sub-populations. We highlight patients with polycystic kidney disease (PKD), as shown in Figure 3. We see that a large proportion of PKD patients are overlapping the Group 3 cluster enriched for heart disease codes. This is consistent with literature reporting PKD patients to be at increased risk for cardiac complications(Ecder 2013). Another cluster of PKD patients visible from Figure 3 can be found in the lower right quadrant of the UMAP plot. We can compare these two regions of enriched PKD patients, examining terms which are under or over-enriched, as shown in Figure 4. Terms which are differentially enriched between the two clusters characterize the differences between the two groups.

**Figure 3:**
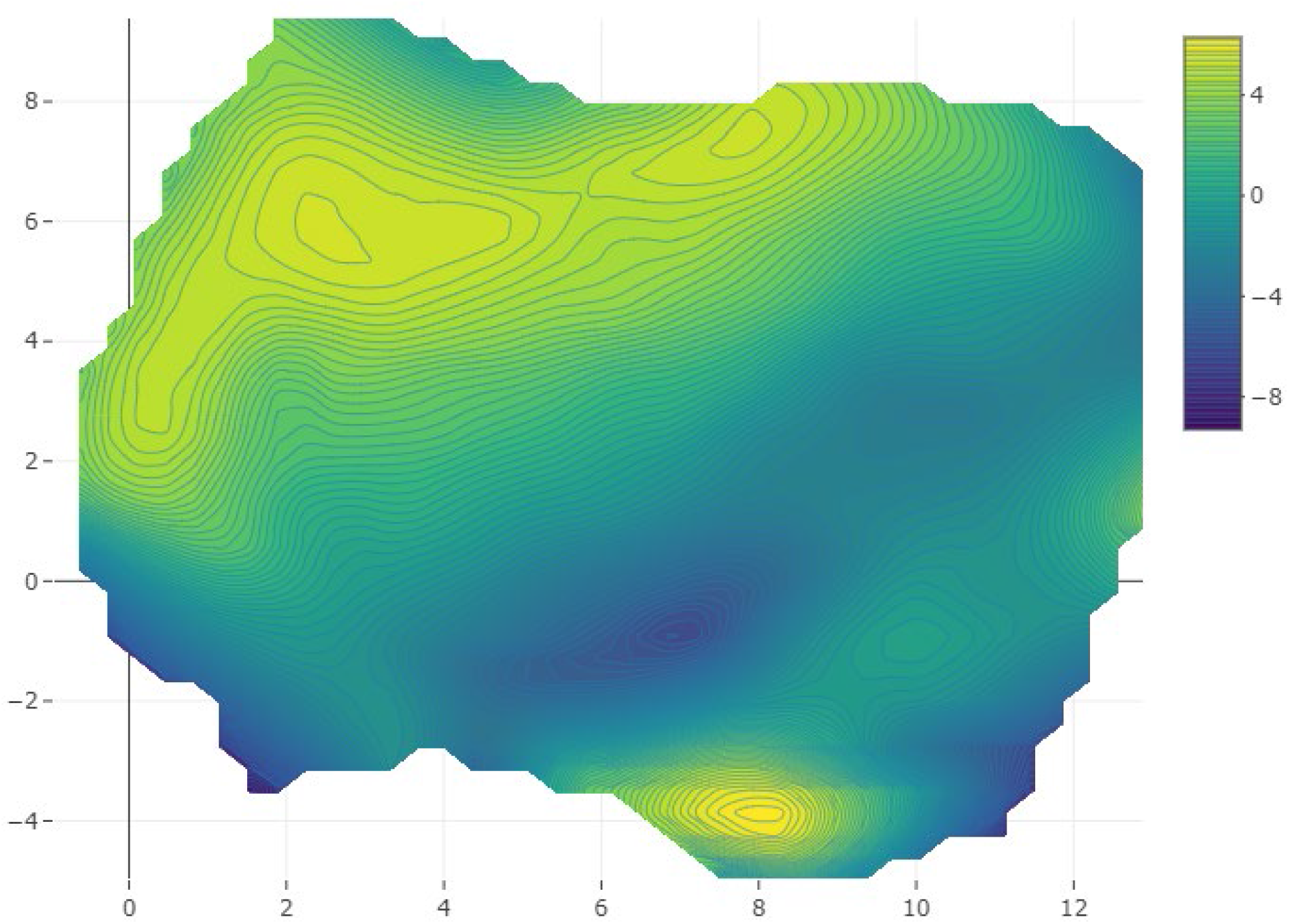
Displaying enrichment of Polycystic kidney disease patients with respect to the UMAP projections of patient-level embeddings.

**Figure 4:**
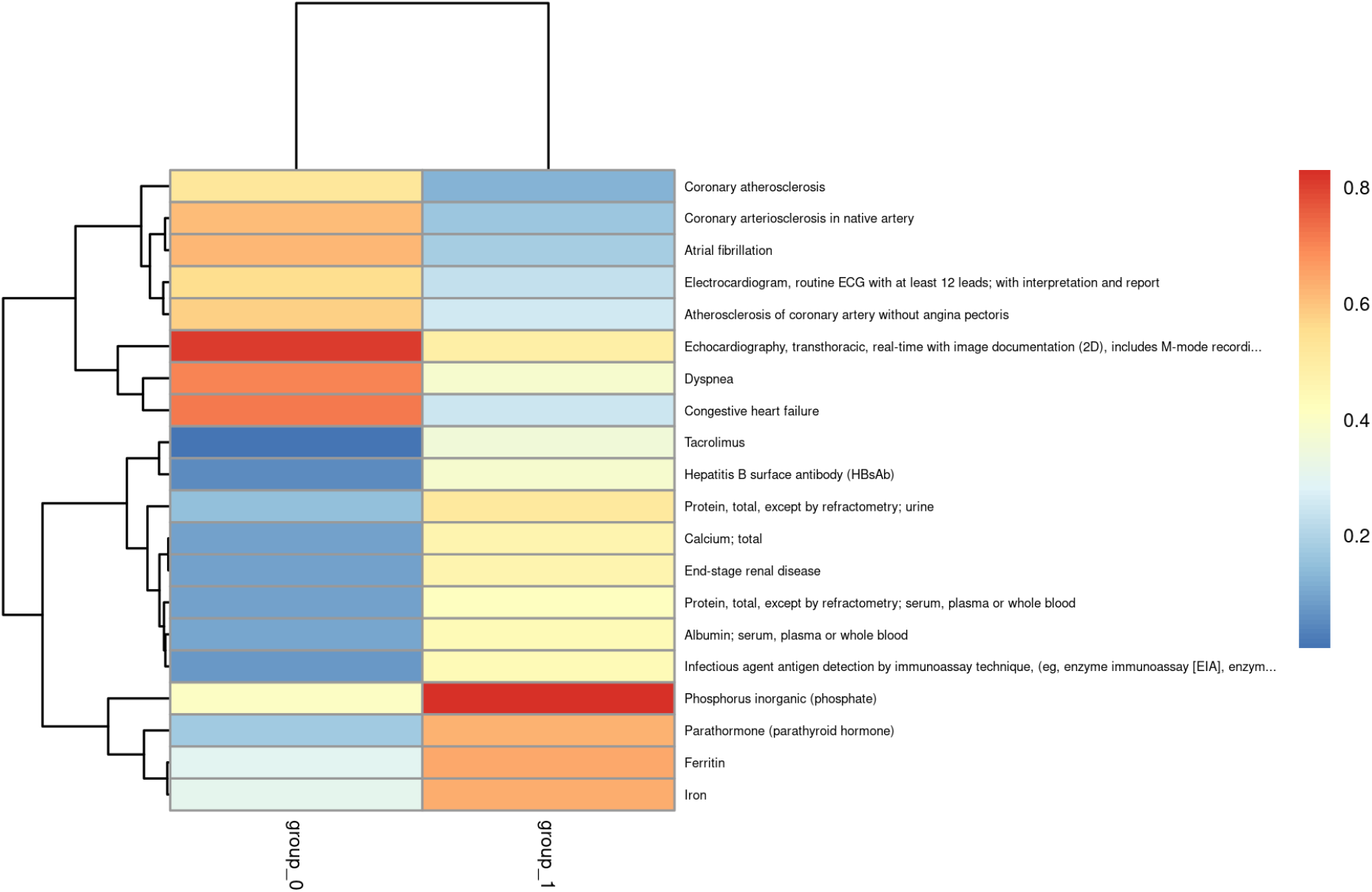
Heatmap displaying the proportion of patients with a given code. The top 20 codes with largest absolute differences between the two regions enriched for PKD patients are shown here. Group 0 refers to patients belonging to the upper cluster and Group 1 refers to patients belonging to the lower cluster seen in Figure 3.

We additionally characterized the population in terms of disease progression to examine which patients underwent rapid progression to end stage renal disease (ESRD) within a given timeframe. We used disease progression quantitative labels, representing the probability of a patient experiencing end stage renal disease (ESRD) within a given timeframe. Such labels were derived by adapting a validated CKD staging electronic phenotype (Shang et al. 2021) which derived a combination of rules based on laboratory measurements, diagnosis and procedure codes to determine a patient’s CKD disease stage (see Methods for further details). Using the probability of progressing from CKD stage 4 to kidney failure within 5 years, we identify groups of patients who rapidly progress to ESRD, as seen in Figure 4. Selecting a sub-population enriched in fast progression to ESRD from the bottom portion of the UMAP plot, we examine terms which are under or over-represented, comparing the region of interest to the complement, as shown in Figure 5. These enriched terms represent hypotheses for factors associated with rapid CKD progressors.

**Figure 5:**
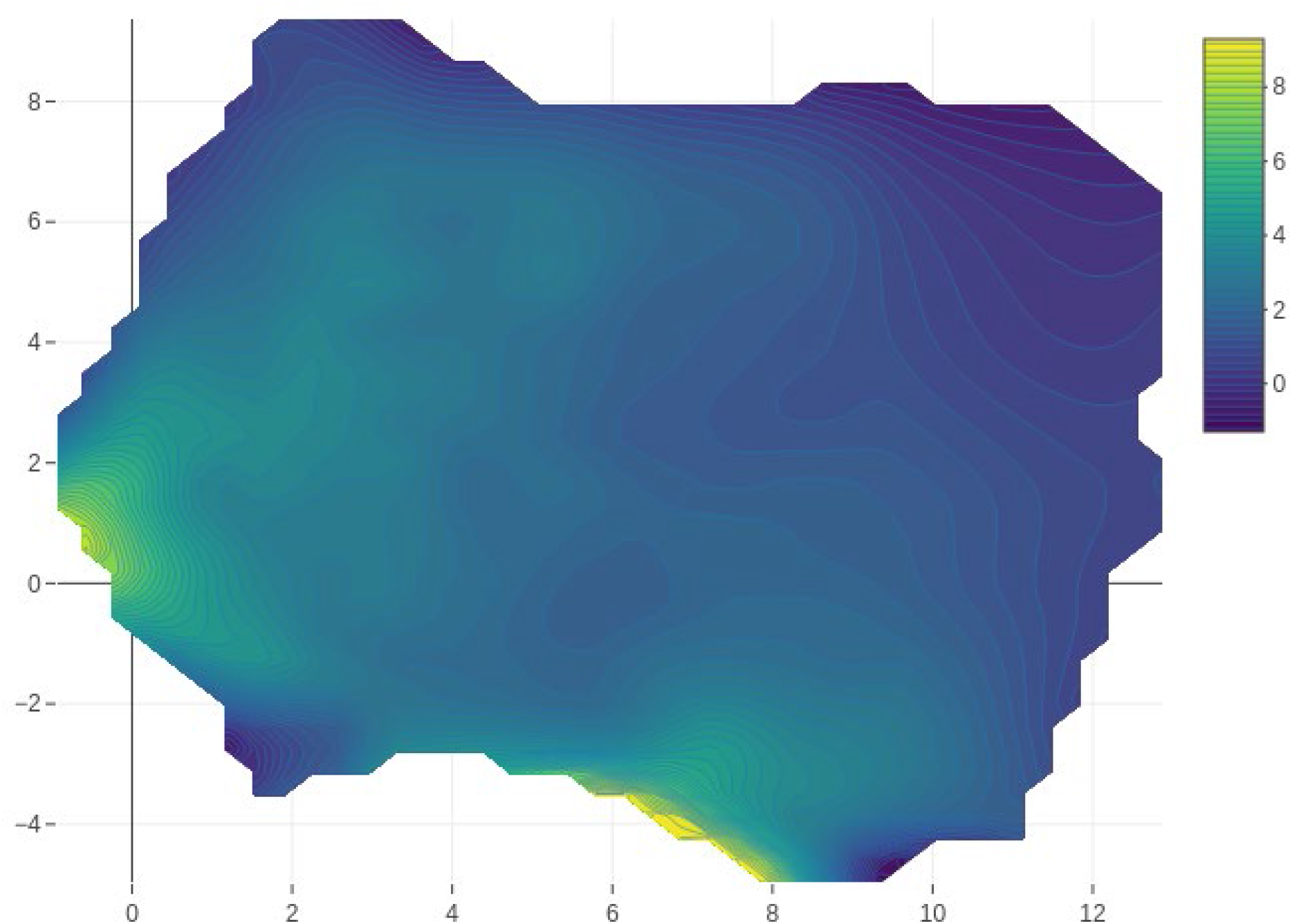
Displaying the logit probability of progressing from CKD stage 4 to kidney failure within 5 years with respect to the UMAP projections of patient-level embeddings.

**Figure 6:**
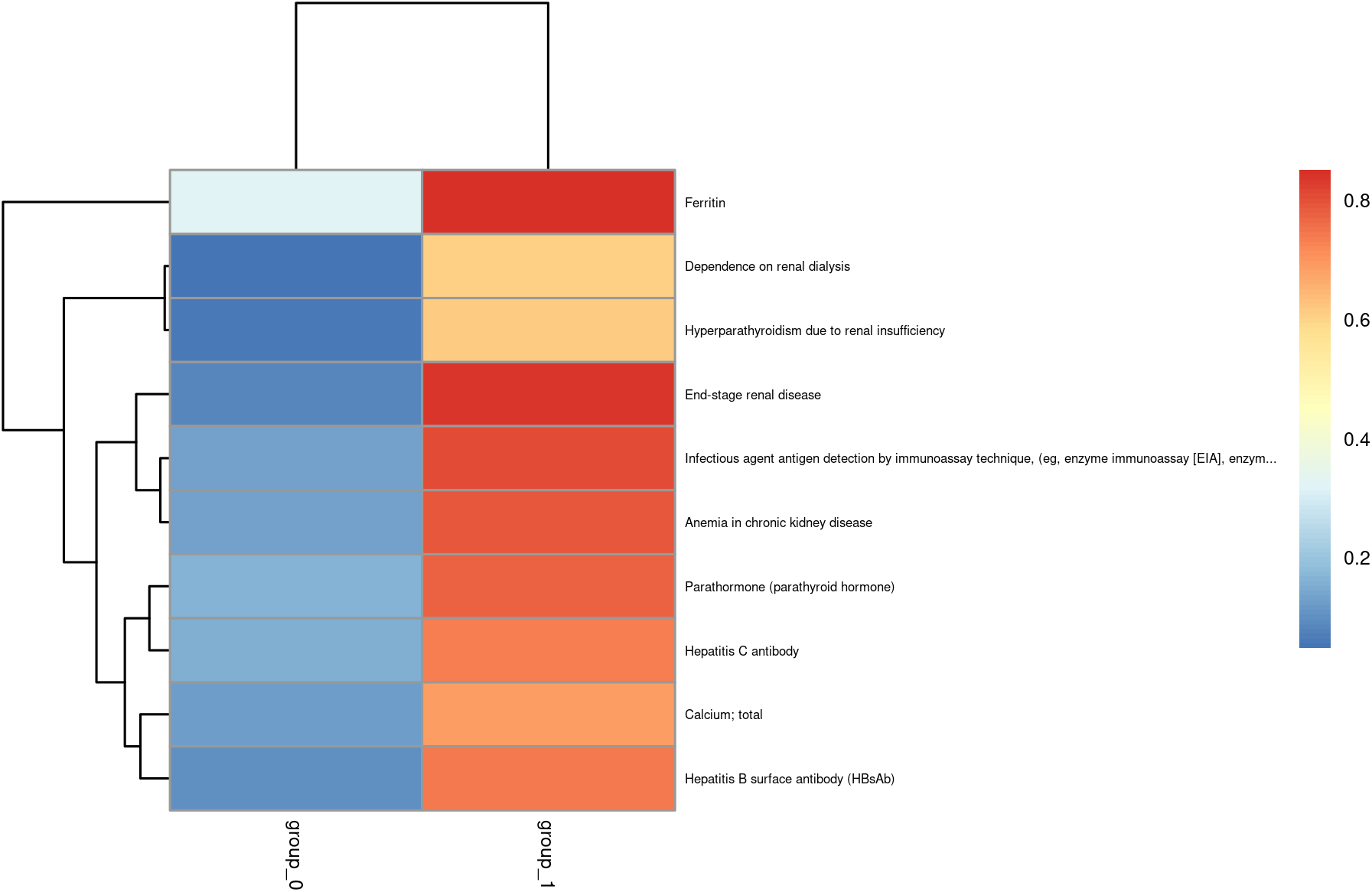
Heatmap displaying the proportion of patients with a given code. The top 10 codes with largest absolute differences between the fast kidney failure progressors compared to the complement are shown here.

## Methods

Optum® de-identified Electronic Health Record dataset, from a US-based patient population, was used, which combines data from a variety of healthcare organizations. We selected patients in the integrated delivery network (IDN) to mitigate against missing data, with the assumption that most healthcare interactions would be met within the network and therefore captured in the database.

To select a kidney disease cohort, we selected all patients with at least one of the diagnosis or procedure codes listed in Supplementary Table 1. We required a minimal ≥ 2 year follow-up period and a minimal number (≥ 15) of serum creatinine measurements.

For each patient in the kidney disease cohort, we extracted diagnosis and procedure codes. ICD9/ICD10 diagnosis codes were mapped to Snomed when possible and otherwise the original codes were retained. CPT4 procedure codes were used. Diagnosis and procedure codes were preprocessed using the same approach described in (Landi et al. 2020b). Briefly, the filtering score is the product of two terms: (i) the proportion of patients with at least one occurrence of a given code and (ii) number of occurrences of a given code in a patient’s trajectory divided by the total number of codes for that patient, summed across all patients. All diagnosis and procedure codes with a filtering score below 1e-8 were excluded. For each patient’s sequence of medical codes, we removed duplicates and performed random shuffling within time windows of 15 days. Patient sequences of medical codes were subsequently chopped into subsequences of fixed length L = 32 to train the ConvAE model.

ConvAE was trained on pre-processed diagnosis and procedure codes. ConvAE was trained on 60% of the data, validated using 20% of the data and a remaining 20% was withheld for testing purposes. The optimal number of training epochs was determined by minimizing the test set cross entropy loss. ConvAE fits an autoencoder which learns a lower-dimensional numerical representation from the input of sequential medical codes. Consequently, for each patient, the learned 100-dimensional embedding was extracted given the sequence of medical codes. When patients had multiple 32-length feature sequences, the average was used.

In order to cluster patients, the 100-dimensional embeddings were projected to 10 dimensions using UMAP, a scalable non-linear dimension reduction technique which has been shown to preserve the global structure of the data (McInnes, Healy, and Melville 2018). K-means clustering was used to determine the clusters. For visualization purposes, the 100-dimensional embeddings were projected to 2 dimensions using UMAP. Code enrichments were considered for codes with frequency at least 1%. CKD disease progression was determined by adapting a validated CKD staging electronic phenotype (Shang et al. 2021) which derived a combination of rules based on laboratory measurements, diagnosis and procedure codes to determine a patient’s CKD disease stage. Whereas the published CKD staging electronic phenotype classifies a patient’s CKD disease stage at the most recent timepoint, we adapted this algorithm to derive a CKD disease stage at all timepoints.

We used eGFR measurements calculated using CKD-EPI formula. As in (Shang et al. 2021), we used their annotated set of codes to distinguish abnormal kidney function that is caused by acute kidney injury or other acute conditions from abnormal kidney function due to chronic conditions. eGFR values co-occurring with such acute events were therefore masked for chronic kidney disease trajectory modeling. A loess curve was fit to the remaining eGFR values that did not co-occur with specific acute events. For each patient and for each day, we classified the CKD stage based on the loess predicted eGFR value and other clinical information to date. We identified the first day of a CKD stage in a monotonic fashion. For example, if a patient was classified as having CKD stage 4 on a given date, and classified as having CKD stage 3 on a later date, the latter CKD stage 3 was ignored and it was assumed that the patient had CKD stage 3 at some timepoint prior to CKD stage 4. We examined the time from a patient’s first occurrence of a given CKD stage to end stage renal disease. When no end stage renal disease was observed, the time was considered censored.

To correlate embedding visualizations with variables of interest (PKD or the probability of ESRD within a timeframe), we performed loess smoothing using the logit probabilities. Regions of interest were defined in a heuristic way. For the subpopulation of PKD patients, we selected all patients corresponding to regions of the UMAP plot where logit probability > 4, either in the upper or lower portion of the UMAP plot. For the subpopulation of patients rapidly progressing to ESRD, we selected all patients in the lower portion of the UMAP plot where logit probability > 8.

## Discussion

This study presents an approach to understand a chronic kidney disease population and generate hypotheses about disease sub-types. It applies a machine learning approach to derive numeric patient representations from sequences of clinical patterns in electronic health records. We propose a visual, exploratory data-driven approach to understand heterogeneity in CKD and to identify potential factors leading to rapid progression of CKD to ESRD. This exploratory approach identifies a high-risk patient population that could be targeted for early intervention to prevent loss of kidney function. The development and application of algorithms using vast electronic health record datasets have the potential to yield data-driven insights into disease populations.

Machine learning approaches together with algorithms relying on expert clinical knowledge offer a powerful approach to define and explore phenotypes. Machine learning algorithms are capable of handling a large number of features and can identify complex patterns in the data. Although the lack of labeled data in observational EHR databases is a significant challenge, we can take advantage of the vast amount of patient-level data to aggregate signals. We took advantage of both expert knowledge used to define specific phenotype labels as well as the large population size, over which we averaged the signals. The use of unsupervised methods rather than supervised methods, which focus on a specific prediction task, has the promise to capture a broad set of information that can be used to understand various aspects of disease.

There are several limitations to our approach. Identifying patients from EHRs is susceptible to selection biases. Only those patients with sufficient data collected in the EHR database were included in our study. Another limitation is that the identified potential risk factors are selected regardless of the timing of the event. Confirmatory studies would be needed to test whether a factor is predictive of the outcome of interest.

We envision several future advancements that could aid in understanding EHR sub-populations. The development of analysis frameworks that combine both expert curated phenotypes and machine learning methods would take advantage of existing knowledge as well as the data-driven learning approaches. The ConvAE unsupervised patient representation learning approach can be compared with other approaches, such as self-supervised approaches which use pseudo-tasks such as masking and predicting information (Devlin et al. 2018) from which numeric representations could also be derived. Alternatively, supervised or weakly supervised approaches could also result in useful learned patient-level representation signals. Having accessible datasets annotated by disease subtypes would greatly aid in the development and evaluation of such methods.

The development of models which can flexibly handle longitudinal data and can incorporate a variety of data types, such as biomarkers, genetics or gene expression may generate more in-depth understanding of individual differences and ultimately enable personalized medicine. Widespread adoption of EHRs would aid in having disease populations from multiple geographies represented, ultimately leading to better insights into diseases.

## Supporting information

coi_disclosure

## Data Availability

All data produced in the present work are contained in the manuscript

## Acknowledgements

We would like to thank Sebastian Hoersch and Rima Izim for helpful comments on the manuscript.

## Supplementary figures

**Supplementary figure 1:**
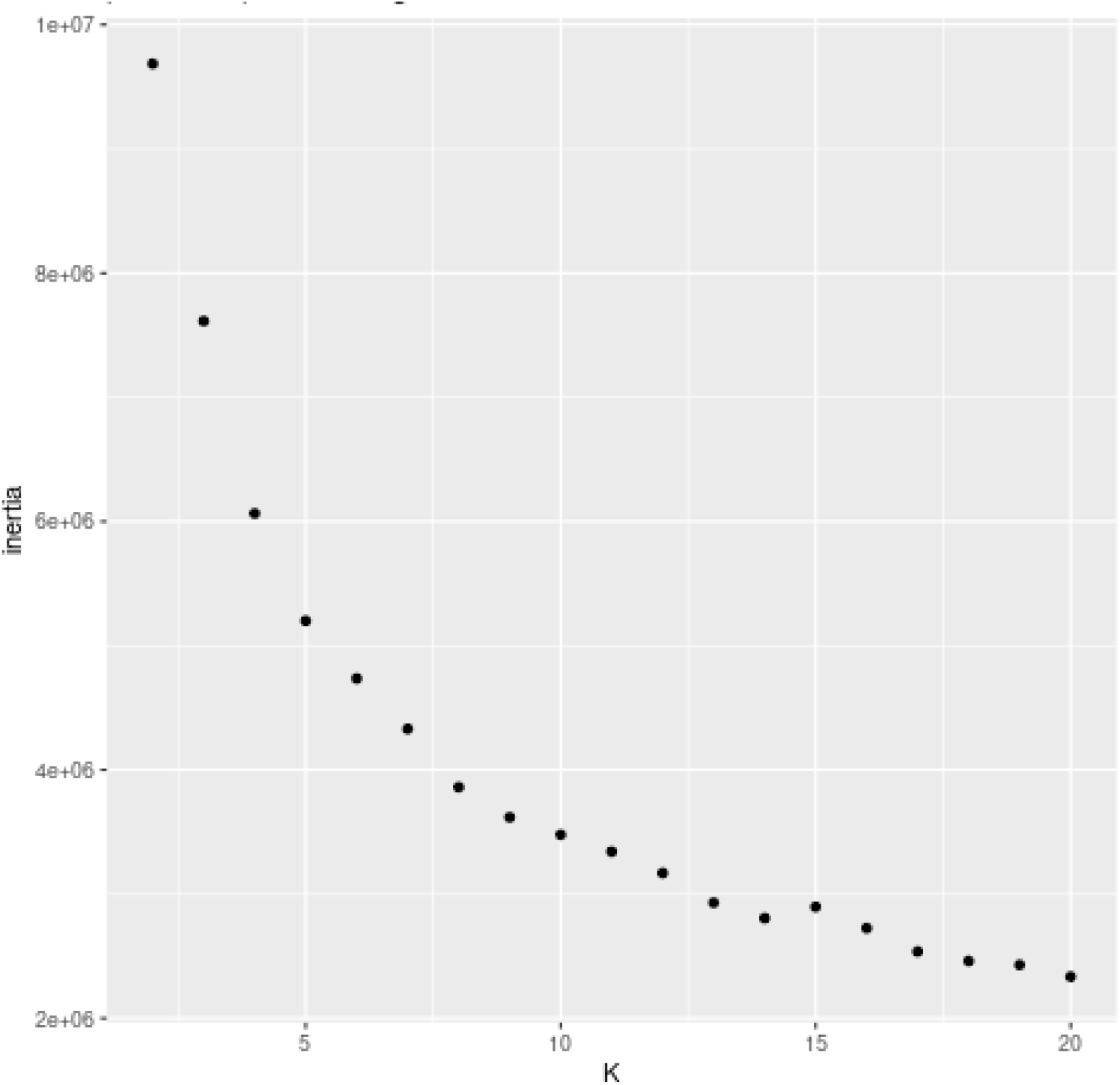
Elbow plot

### Supplementary tables

Supplementary Table 1: Kidney disease diagnosis/procedure codes used for selecting the patient cohort. The set of codes consists of diagnosis codes relating to prerenal injury, chronic kidney disease, acute kidney injury, dialysis, kidney transplant or other kidney disease and procedure codes relating to dialysis or kidney transplant published in (Shang et al. 2021).

Supplementary Table 2: The proportion of patients with a given code for each of the K=5 clusters.

